# Shared genetic features between lymphoma pathogenesis and oestrogen levels

**DOI:** 10.1101/2024.05.17.24307324

**Authors:** Claire Hill, Marisa Muckian, Marisa Cañadas Garre, Graeme Greenfield, Alexander P. Maxwell, Charlene M. McShane, Lesley Ann Anderson, Amy Jayne McKnight

## Abstract

In the last decade, genetic analysis has aided lymphoma classification, alongside traditional phenotypic subdivision. Lymphomas exhibit sex differences in disease incidence and prevalence; therefore, scope existed to explore sex-specific genetic variation.

Harnessing UK Biobank genetic data (185,000 females, 176,000 males), sex-specific genome-wide genetic variation significantly associated with lymphoma subtypes was determined in both autosomal and sex chromosome genes. Sex-specific genetic variation significantly associated with sex-related characteristics was also identified. Functional predictions for those genes containing significant (p<5×10^-5^) genetic variation was carried out via ViSEAGO. For each lymphoma subtype, approximately 10-12% of the genes containing significantly associated genetic variants were shared between the sexes, highlighting largely sex-specific profiles for lymphoma subtypes.

Significantly associated X chromosome genetic variants were identified for Non-Hodgkin’s lymphoma (NHL) and Diffuse large B-cell lymphoma (DLBCL), such as variants within *SYTL5* (in males) and *PPP2R3B* (in females); genes previously implicated in haematological malignancy biological processes. Additionally, in female NHL patients, a genetic variant mapped to *ESR1*, the gene coding for oestrogen receptor-α was identified, adding to the body of evidence highlighting the relevance of oestrogen regulation for haematological malignancy subtypes.

Indeed, up to 9.1% of overlap was observed between those genes containing significant genetic variation associated with specific lymphoma subtypes, and those significantly associated with oestrogen level determination. Gene ontology analysis further emphasised functional overlaps between lymphoma and oestrogen level determination in females, highlighting predicted involvement in epigenetic modification, gene expression, and signalling.

This study revealed sex-specific genetic variation significantly associated with lymphoma, as well as hormone level determination, and revealed biological pathways potentially disrupted during lymphoma pathogenesis which are influenced by oestrogen. Exploring these pathways may advance our understanding of the sex-specificities of lymphoma, and may reveal therapeutic targets to advance patient care.

## Introduction

In recent decades, the understanding of haematological malignancies, including lymphoma, has rapidly evolved, partly due to the integration of genomic analyses facilitating the discovery of both similarities and differences between disease subtypes [1,2]. Lymphomas can be broadly categorised into Hodgkin’s lymphoma (HL) and Non-Hodgkin’s lymphoma (NHL). NHL can be further sub-categorised into B-cell subtypes, such as diffuse large B-cell lymphoma (DLBCL) and follicular lymphoma (FL), or T-cell subtypes [3,4]. Scope remains to further explore the genetic profile of lymphoma and its subtypes, particularly in relation to sex differences, to advance diagnosis and aid treatment planning.

Incidence of cancer is often higher in males [5–8]. Data collected in a UK population between 2010 to 2016 determined that the male to female sex-rate ratio for all haematological malignancies was 1.37 [1,9]. Globally in 2019, NHL and HL were 1.57 and 1.29 times more prevalent in males compared to females, respectively [10]. In the same year, deaths caused by NHL and HL were 1.34 and 1.65 times higher in males compared to females globally [10]. Sex differences in processes such as hormone-mediated regulation, viral infection, immunity, gene expression, stress response and/or homeostatic cellular functions may contribute to variations in incidence and/or prevalence rates between sexes [11–13].

Several genetic mediators may be responsible for the sex differences observed in lymphoma incidence. Sex chromosomes and sex hormones are intricately linked in immunoregulatory processes [14]; affecting autoimmune disease susceptibility [15,16] and progression of haematological malignancies [17]. Distinct differences exist between immune function in males and females, with influence from genetic factors [18]; however, sex differences with respect to disease incidence and progression are not fully understood. Scope existed to explore the genetic landscape of lymphoma to determine the influence of sex and sex-related characteristics [i.e. circulating serum hormone levels, including oestrogen, testosterone and sex hormone binding globulin (SHBG)] on these associations, in a UK Biobank (UKB) cohort.

## Methods

### Study Design

This project harnessed a genome-wide association (GWA) approach, analysing single nucleotide polymorphisms (SNPs) across all chromosomes to investigate their association with lymphoma and serum hormone levels in a population cohort. The analyses were stratified by biological sex and lymphoma subtype.

### Participants

The cohort consisted of UKB participants up until 08/2019 [19]. The UKB is a long-term United Kingdom-based study that aims to research the impact of genetic and environmental contributions to the predisposition of disease. Participants, aged 40-69 years old, were recruited between 2006 and 2010. Only individuals with a Caucasian ethnic background, the predominant ethnicity in the UKB, as genetically determined by the UKB [20], were included in this study. These selected individuals have a very similar ancestral background relative to the full UKB cohort [20]. In the UKB, 86% of individuals identified themselves as ‘White British,’ with the Caucasian ethnic grouping defined by the UKB as individuals who self-identified as ’White British’ and have very similar genetic ancestry based on a principal components analysis of the genotypes. Women with hysterectomy were excluded.

### Outcome variables

#### Cancer outcomes

To identify patients with lymphoma, incident and prevalent cases were considered, and defined as per the ‘Type of Cancer ICD9/ICD10’. Relevant information about the ICD codes used to define lymphoma in this study, and distinct subtypes, can be found in S1.

#### Serum variables

Serum hormone measurements of oestrogen, testosterone and sex hormone binding globulin (SHBG), respectively, were analysed in this study.

#### Clinical and sociodemographic variables

The clinical and sociodemographic variables included body fat percentage, body mass index (BMI), education, social economic status, abdominal obesity, alcohol consumption, smoking status and age. Female-specific covariates included age at menarche, menopausal status, age of menopause, current use of oral contraceptives and current use of hormone replacement therapy (HRT). Male specific covariates included relative age of first facial hair, relative age voice broke and hair balding pattern. Relevant information used to define the covariates can be found in the definition of the sociodemographic and clinical variables in S2.

### Genotyping and Quality Control

The Applied Biosystems^TM^ UKB Axiom^TM^ and UK BiLEVE Axiom^TM^ Affymetrix Arrays were used for genotyping. Genotypes for autosomes, X and XY chromosomes were imputed into a reference dataset combining the Haplotype Reference Consortium (HRC) and UK10K haplotype resource [20]. PLINK (v2.0) was used to perform quality control [21]. Individuals with excessive sample heterozygosity (>95%) or missingness (>95%) were removed. Related individuals (identity by kinship coefficient >0.0884) and principal component (PCA) outliers, as calculated by the UKB, were also removed. Other exclusion criteria for samples were discordance between inferred sex and reported sex, kinship not calculated, putative sex chromosome aneuploidy or consent withdrawn, according to the centralized sample quality control performed by UKB [20]. SNPs with an excessive missingness genotyping rate (>95%), imputation quality <0.3, variant call rate <95%, minor allele frequency ≤0.001 or with a minor allele count lower than 5, and those which deviated from the Hardy-Weinberg equilibrium (HWE) (p<1×10^-20^) [22], were removed. We considered a threshold for statistical significance of 5×10^-8^. Clumping for associated SNPs was performed with PLINK-1.9 to identify blocks of variants in linkage disequilibrium with R2 >0.1 and <500KB from the index variant (smallest p-value) [23]. After clumping SNPs were annotated using Anno [24]. The frequency of the effect allele for each variant in the general population was compared to the 1000 genomes project database, Great Britain sub-population [25].

### Statistical Analysis

Descriptive analysis was performed using R [26]. The qualitative variables were notated as a percentage (%) of their total (N). Normality of the quantitative variables was assessed using the Kolmogorov-Smirnov test and non-normally distributed quantitative variables were subsequently expressed by their median and interquartile range. Lymphoma cases versus control association analysis was performed using PLINK (v2.0) for the whole UKB cohort, stratified by sex [21]. Logistic regression was performed for the cancer outcome variables. Control groups for each lymphoma analysis were defined as the entire cohort excluding the case group of interest. The quantitative phenotypes (oestrogen, testosterone and SHBG) were normalised (using natural logarithm of the variable followed by --pheno-quantile-normalize flag in the regression) and linear regression was then performed.

### Gene profile and gene ontology analysis

SNPs identified during each GWA analysis were filtered based on their p-value, with only those with p<5×10^-5^ retained for downstream analysis. rsIDs for each SNP were assigned to nearest genes using the online Variant Effect Predictor (VEP) tool (using Assembly GRCh37, and both Ensembl and RefSeq transcript databases) [27], to generate gene profiles for each lymphoma and hormone grouping. Gene ontology analysis of biological processes associated with each gene profile, as well as the subsequent clustering of gene ontology terms, was performed using the ViSEAGO package (v. 1.6.0), harnessing the TopGO package (v. 2.44.0), in R studio (v. 1.4.1717), R version 4.1.1.[28–31] The background gene list of all human genes was generated using the R package, org.Hs.eg.db (v. 3.12.0) [32]. As necessary, genes were converted to their relevant Enterez IDs using the org.Hs.eg.db database in R studio, with retired Ensembl IDs excluded from downstream analysis. ViSEAGO Enterez annotations (ID: 9606) were harnessed. Fisher’s exact test, with either the *classic* or *elim* algorithm, was used to identify enriched gene ontology terms, utilising a significance cut off of p<0.01.

## Results

Harnessing the UKB genetic variant data, 14,015,115 variants were available for analysis after quality control processes (S3). Analysis of the UKB cohort was stratified by sex, and after quality control, 184,843 females and 176,399 males remained (Fig 1). The descriptive analysis of each lymphoma and hormone grouping, along with the sociodemographic variables of these UKB participants, is summarised in Table 1. Across all lymphomas described in Table 1, a higher percentage of cases was observed in males, compared to females.

**Fig 1.**
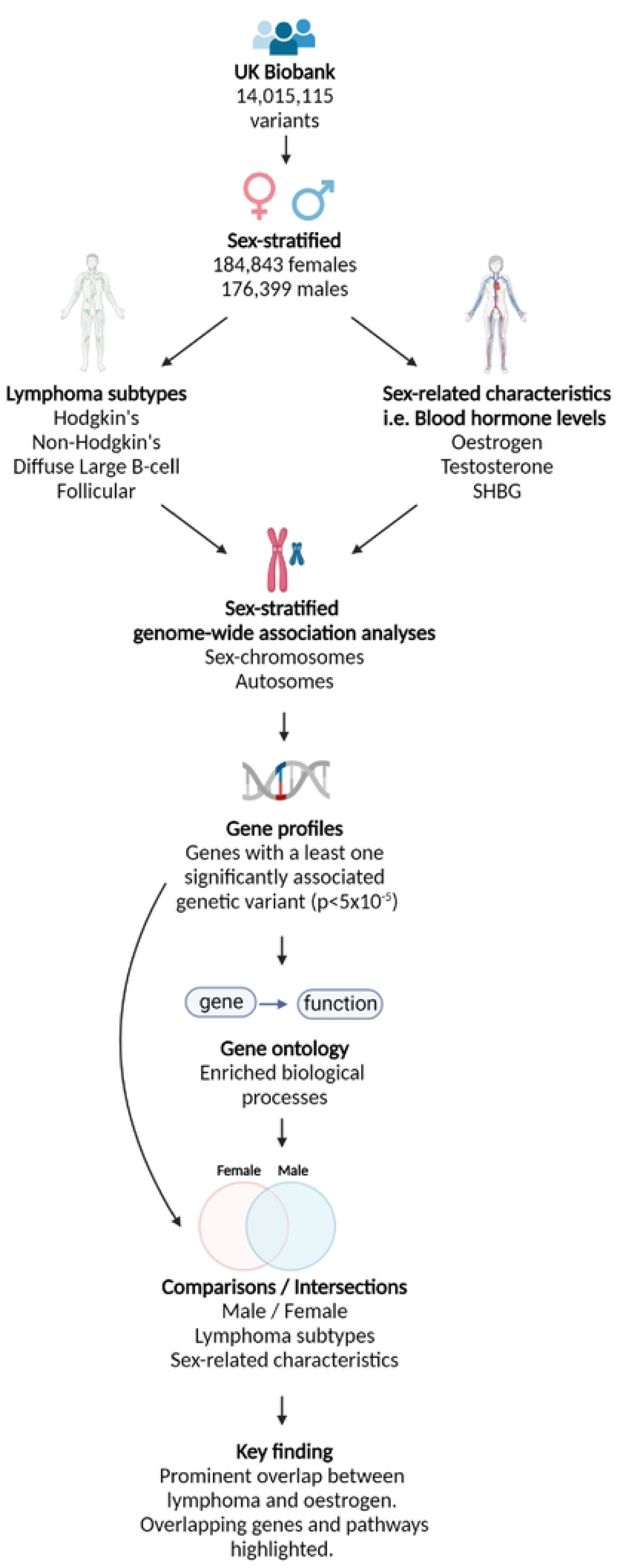
Flow diagram representing the data analysis pipeline. First stratifying the UK Biobank cohort into males and females, followed by genome-wide association (GWA) analyses to determine genetic variants significantly associated with lymphoma outcomes (Hodgkin’s lymphoma (HL), Non-Hodgkin’s lymphoma (NHL), Diffuse large B-cell lymphoma (DLBCL) and Follicular lymphoma (FL)), or sex-related characteristics (testosterone, oestrogen or SHBG hormone levels). Gene profiles were generated for each lymphoma outcome and sex-related characteristic by selecting those genes presenting at least one significantly associated genetic variant (p<5×10^-5^). Gene ontology analysis was carried out on these gene profiles to determine enriched biological processes associated with lymphoma subtypes or sex-related characteristics. These gene and gene ontology profiles could be compared between males and females, lymphoma subtypes and sex-related characteristics to highlight similarities and differences between these groupings, highlighting novel overlapping and specific pathways useful for the development of future studies to as they hold potential to be associated with sex-specificities of lymphoma pathologies.

**Table 1.** Descriptive analysis of patients from the UK Biobank Cohort. Patients have been stratified by sex and the variables are described separately for females and males. BMI=Body mass index, OC=Oral contraceptive, HRT=Hormone replacement therapy.

|  | Females |  | Males |  |
| --- | --- | --- | --- | --- |
|  | N | (%) P50[P25-P75] | N | (%) P50[P25-P75] |
| Age | 184843 | 57[50-63] | 176399 | 59[51-64] |
| Alcohol Consumption (Units per Week)<br>(≤4.66/>4.66-≤14/>14/Former Drinker) | 184694 | (12.1/58.9/25.5/3.5) | 176219 | (5.6/37/54.2/3.2) |
| Smoking Status<br>Never/Previous/Currently | 184227 | (59.8/31.6/8.6) | 175775 | (48.9/39.3/11.8) |
| **BMI**<br>**≤24.99/≥25-≤29.99/≥30** | 183920 | (40.7/36.8/22.5) | 175424 | (24.9/49.7/25.4) |
| **Body Fat Percentage**<br>**P[25-100]** | 181918 | (6.7/16.1/34.9/43.3) | 172915 | (49/36.2/13.2/1.6) |
| **Education**<br>**None/GCSE(or equivalent)/A-Level(or equivalent)/Degree** | 183418 | (16.9/20.4/32.1/30.6) | 174850 | (17.7/13.9/35.7/32.7) |
| **Socioeconomic Status**<br>**Affluent/Semi-Affluent/Middle/Semi-Deprived/Deprived** | 184639 | (21.5/21.3/21.1/19.7/16.4) | 176187 | (21.6/21.2/20.5/19.2/ 17.5) |
| **Abdominal Obesity** | 184524 | (18.3/25.3/26.7/18.2/11.5) | 176089 | (29.1/31/26.2/9.3/4.4) |
| **Age Menarche**<br>**<13/13-14/≥15** | 179836 | (38.8/44.7/16.5) | - | - |
| **Menopausal Status**<br>**Pre/Post** | 163260 | (27.9/72.1) | - | - |
| **Age Menopause**<br>**<50/≥50** | 110248 | (35.2/64.8) | - | - |
| **Current OC use**<br>**Yes/No** | 184,843 | (1.9/98.1) | - | - |
| **Current HRT use**<br>**Yes/No** | 184,843 | (5/95) | - | - |
| **OC use**<br>**<5 years/ ≥ 5 years** | 127,027 | (22.2/77.8) | - | - |
| **HRT duration**<br>**<5 years/ ≥ 5 years** | 41397 | (35.3/64.4) | - | - |
| **Relative Age Voice Broke**<br>**Younger than average/Average/Older than average** | - | - | 163572 | (4.2/89.9/5.9) |
| **Relative Age of First Facial Hair**<br>**Younger than average/Average/Older than average** | - | - | 170904 | (6.4/80.6/13) |
| **Hair Balding**<br>**Yes/No** | - | - | 175252 | (68.2/31.8) |
| **All Lymphoma Cases** | 778 | (0.42) | 1152 | (0.65) |
| **All Non-Hodgkin's Lymphoma** | 633 | (0.34) | 969 | (0.55) |
| **All Hodgkin's Lymphoma** | 154 | (0.08) | 200 | (0.11) |
| **T Cell Lymphoma** | 48 | (0.03) | 89 | (0.05) |
| **DLBCL** | 186 | (0.1) | 306 | (0.17) |
| **Follicular Lymphoma** | 164 | (0.09) | 205 | (0.12) |
| **Oestrogen** | 42833 | 400.4[266.7-639.5] | 14975 | 203.9[188.9-231] |
| **Testosterone** | 158990 | 1.025[0.731-1.386] | 175461 | 11.612[9.421-14.116] |
| **SHBG** | 42833 | 56.87[40.55-77.4] | 162331 | 37.34[28.34-48.6] |

To further explore these sex differences, the genetic variation of those with different lymphoma subtypes was investigated via a series of GWA analyses, in males and females separately, as outlined in Fig 1. The top five most significant associations for each lymphoma subtype are summarised in Table **2** (autosomal) **Table 2** and Table 3 (sex chromosome encoded). Literature detailing previous associations between these autosomal or sex chromosome genes and the development or progression of haematological malignancies has been summarised in S4 and S5.

**Table 2.** Top five SNPs associated with lymphoma subtypes, separated into male and female cohorts. Abbreviations: A1: affect allele, Alt: alternate allele, Chr: chromosome, Ref: reference allele, SE standard error, SNP: single nucleotide polymorphism.

| Lymphoma grouping | Sex | Gene | Index SNP (Total Linked) | Chr | Position | Ref | Alt | A1 | Functional Consequence | OR | SE | P-Value |
| --- | --- | --- | --- | --- | --- | --- | --- | --- | --- | --- | --- | --- |
| Lymphoma | Female | *CRB1* | rs192495315 (0) | 1 | 197178872 | A | G | A | Intron Variant | 24.428 | 0.497098 | 1.29x10<sup>-10</sup> |
|  |  | *GRIK4* | rs79342674 (2) | 11 | 120481542 | G | C | G | Intron Variant | 3.28294 | 0.18937 | 3.44x10<sup>-10</sup> |
|  |  | *XKR9* | rs55830581 (4) | 8 | 71867044 | A | G | A | Intron Variant | 8.77028 | 0.347533 | 4.16x10<sup>-10</sup> |
|  |  | *ALCAM* | rs181809302 (0) | 3 | 105169099 | C | T | C | Structural Variation (Intron) | 22.3496 | 0.497364 | 4.20x10<sup>-10</sup> |
|  |  | *BIRC6* | rs750820813 (1) | 2 | 32727344 | G | A | G | Structural Variation (Intron) | 35.8666 | 0.574271 | 4.56x10<sup>-10</sup> |
|  | Male | *MARVELD2* | 5:68724317_A_G (9) | 5 | 68724317 | G | A | G | Structural Variation (Intron) | 23.0065 | 0.494656 | 2.31x10<sup>-10</sup> |
|  |  | *SLC25A15* | rs574707243 (1) | 13 | 41364460 | T | G | T | Structural Variation (Intron) | 7.25363 | 0.332854 | 2.63x10<sup>-09</sup> |
|  |  | *NCALD* | rs117349373 (7) | 8 | 102930704 | T | G | T | Structural Variation (Intron) | 3.20388 | 0.19678 | 3.28x10<sup>-09</sup> |
|  |  | *ARFIP2* | rs190414132 (1) | 11 | 6502011 | C | G | C | Structural Variation (Intron) | 6.85473 | 0.328862 | 4.82x10<sup>-09</sup> |
|  |  | *PTPRD* | 9:8980401_CAG_C (2) | 9 | 8980401 | C | CAG | C | Structural Variation (Intron) | 5.81783 | 0.301211 | 5.03x10<sup>-09</sup> |
| Hodgkin's lymphoma | Female | *GTF2IP23* | rs548617742 (10) | 7 | 66326622 | A | G | A | Intron Variant | 82.7804 | 0.65199 | 1.26x10<sup>-11</sup> |
|  |  | *ZNF273* | rs553732942 (4) | 7 | 64379561 | C | T | C | Structural Variation (Intron) | 148.026 | 0.742687 | 1.71x10<sup>-11</sup> |
|  |  | *UGT2B15* | rs543475625 (6) | 4 | 69410572 | G | A | G | Structural Variation (Intron) | 136.523 | 0.732694 | 1.94x10<sup>-11</sup> |
|  |  | - | rs140400666 (2) | 7 | 72835692 | A | G | A | Intergenic | 65.8162 | 0.624046 | 1.96x10<sup>-11</sup> |
|  |  | - | rs545988972 (4) | 17 | 36837313 | G | A | G | Intergenic | 116.402 | 0.70951 | 2.02x10<sup>-11</sup> |
|  | Male | *DTNB* | rs535941145 (0) | 2 | 25755947 | A | C | A | Structural Variation (Intron) | 23.9377 | 0.460015 | 5.09x10<sup>-12</sup> |
|  |  | *LOC105375911* | rs117832099 (2) | 8 | 79333688 | T | G | T | Intron Variant | 17.2194 | 0.424084 | 1.93x10<sup>-11</sup> |
|  |  | - | rs117028336 (1) | 10 | 93478127 | G | A | G | Intergenic | 65.6625 | 0.624405 | 2.06x10<sup>-11</sup> |
|  |  | *PNPLA7* | 9:140367191_G_A (13) | 9 | 140367191 | A | G | A | Structural Variation (Intron) | 25.9762 | 0.493028 | 3.94x10<sup>-11</sup> |
|  |  | - | rs7239000 (13) | 18 | 68549937 | A | G | A | Intergenic | 75.0017 | 0.654643 | 4.25x10<sup>-11</sup> |
| Non-Hodgkin's lymphoma | Female | *ALCAM* | rs181809302 (0) | 3 | 105169099 | C | T | C | Structural Variation (Intron) | 30.3625 | 0.498413 | 7.48x10<sup>-12</sup> |
|  |  | *BIRC6* | rs750820813 (1) | 2 | 32727344 | G | A | G | Structural Variation (Intron) | 49.056 | 0.578073 | 1.65x10<sup>-11</sup> |
|  |  | *ESR1* | 6:152322665_GT_G (4) | 6 | 152322665 | G | GT | G | Structural Variation (Intron) | 28.5795 | 0.502897 | 2.62x10<sup>-11</sup> |
|  |  | *DLGAPS* | rs148504179 (5) | 14 | 55640860 | T | C | T | Structural Variation (Intron) | 14.6337 | 0.40277 | 2.70x10<sup>-11</sup> |
|  |  | - | rs532735679 (6) | 7 | 13134144 | G | T | G | Intergenic | 21.3054 | 0.464419 | 4.50x10<sup>-11</sup> |
|  | Male | *MARVELD2* | 5:68724317_A_G (9) | 5 | 68724317 | G | A | G | Structural Variation (Intron) | 29.4908 | 0.494727 | 7.90x10<sup>-12</sup> |
|  |  | - | rs545246664 (1) | 11 | 76285314 | T | G | T | Intergenic | 4.7031 | 0.245366 | 2.79x10<sup>-10</sup> |
|  |  | *LOC105370932* | rs3849231 (8) | 15 | 83959560 | A | C | A | Intron Variant | 5.48678 | 0.285328 | 2.43x10<sup>-09</sup> |
|  |  | *SLC25A15* | rs574707243 (1) | 13 | 41364460 | T | G | T | Structural Variation (Intron) | 7.92141 | 0.35067 | 3.60x10<sup>-09</sup> |
|  |  | - | rs75350437 (44) | 2 | 138582908 | T | C | T | Intergenic | 48.8383 | 0.662339 | 4.33x10<sup>-09</sup> |
| Diffuse large B-cell lymphoma | Female | - | 8:125780384_TA_T (8) | 8 | 125780384 | T | TA | T | Intergenic | 206.189 | 0.768153 | 4.00x10<sup>-12</sup> |
|  |  | - | rs74134778 (5) | 10 | 55520836 | A | G | A | Intergenic | 73.1804 | 0.632743 | 1.16x10<sup>-11</sup> |
|  |  | *LOC440900* | rs539342948 (9) | 2 | 114753560 | T | C | T | Structural Variation | 108.868 | 0.698552 | 1.89x10<sup>-11</sup> |
|  |  | - | rs557644732 (0) | 15 | 78693186 | C | A | C | Intergenic | 1423.69 | 1.0887 | 2.57x10<sup>-11</sup> |
|  |  | *KCNN2* | rs527342039 (5) | 5 | 113395825 | T | C | T | Intron Variant | 95.4078 | 0.686128 | 3.07x10<sup>-11</sup> |
|  | Male | - | rs76641300 (9) | 8 | 9152845 | T | G | T | Intergenic | 61.1303 | 0.614836 | 2.24x10<sup>-11</sup> |
|  |  | *VIT* | rs112593004 (0) | 2 | 36978664 | A | G | A | Structural Variation (Intron) | 37.845 | 0.548441 | 3.47x10<sup>-11</sup> |
|  |  | *MSI2* | rs527354167 (0) | 17 | 55600653 | A | C | A | Structural Variation (Intron) | 16.62 | 0.424511 | 3.57x10<sup>-11</sup> |
|  |  | *RARB* | rs144624813 (3) | 3 | 25531457 | C | T | C | Structural Variation (Intron) | 6.91193 | 0.292931 | 4.12x10<sup>-11</sup> |
|  |  | - | rs6085653 (0) | 20 | 721111 | G | A | G | Intergenic | 168.5 | 0.783346 | 5.95x10<sup>-11</sup> |
| Follicular lymphoma | Female | - | rs191650498 (0) | 22 | 35919641 | C | G | C | Intergenic | 63.938 | 0.600333 | 4.33x10<sup>-12</sup> |
|  |  | - | rs60792005 (3) | 3 | 94937576 | T | A | T | Intergenic | 96.4836 | 0.676108 | 1.40x10<sup>-11</sup> |
|  |  | - | rs138132722 (3) | 5 | 78477488 | G | C | G | Intergenic | 51.4798 | 0.585766 | 1.72x10<sup>-11</sup> |
|  |  | - | rs534001699 (1) | 3 | 176671111 | A | T | A | Intergenic | 81.7487 | 0.671363 | 5.41x10<sup>-11</sup> |
|  |  | - | rs568377288 (4) | 7 | 13257202 | T | G | T | Intergenic | 77.9685 | 0.665334 | 5.85x10<sup>-11</sup> |
|  | Male | - | rs114814595 (0) | 10 | 104972850 | T | C | T | Intergenic | 29.601 | 0.459855 | 1.74x10<sup>-13</sup> |
|  |  | *SORD2P* | rs551056857 (1) | 15 | 45130497 | T | C | T | Intron Variant | 31.1025 | 0.499043 | 5.67x10<sup>-12</sup> |
|  |  | *NPVF* | rs146193251 (0) | 7 | 25264426 | T | A | T | Structural Variation (Intron) | 227.892 | 0.789239 | 6.04x10<sup>-12</sup> |
|  |  | - | rs531214379 (11) | 5 | 111377236 | T | C | T | Intergenic | 9.59628 | 0.332959 | 1.11x10<sup>-11</sup> |
|  |  | *LOC105377861* | rs79282826 (5) | 6 | 77415091 | A | G | A | Intron Variant | 67.1832 | 0.631257 | 2.64x10<sup>-11</sup> |

**Table 3.** Sex chromosomes SNPs associated with lymphoma subtypes or hormone level determination, stratified by sex. Abbreviations: A1: affect allele, Alt: alternate allele, Chr: chromosome, Ref: reference allele, SE standard error, SNP: single nucleotide polymorphism.

| Sex | Outcome | Gene | Index SNP (Total Linked) | Chr | Position | Ref | Alt | A1 | Functional Consequence | BETA | SE | P-Value |
| --- | --- | --- | --- | --- | --- | --- | --- | --- | --- | --- | --- | --- |
| Female | SHBG | - | rs5942656 (3) | X | 109866545 | G | T | G | - | 0.048989 | 0.00799273 | 8.99x10<sup>-10</sup> |
|  | Lymphoma | - | rs182738927 (0) | XY | 933131 | G | C | G | - | 12.7925 | 0.466644 | 4.71x10<sup>-08</sup> |
|  | Non-Hodgkin's | - | rs181907766 (0) | XY | 932761 | T | C | T | - | 15.8644 | 0.469102 | 3.81x10<sup>-09</sup> |
|  |  | - | rs182738927 (0) | XY | 933131 | G | C | G | - | 16.4072 | 0.474982 | 3.86x10<sup>-09</sup> |
|  | DLBCL | *PPP2R3B* | rs776140450 (0) | XY | 295418 | T | C | T | Intron Variant | 109.697 | 0.798903 | 4.10x10<sup>-09</sup> |
| Male | SHBG | - | rs12013156 (4) | X | 109786110 | A | C | A |  | 0.0954765 | 0.00241566 | P<0.05<sup>-08</sup> |
|  |  | - | rs73251825 (4) | X | 109824249 | G | C | G |  | -0.048746 | 0.0050241 | 2.99x10<sup>-22</sup> |
|  |  | - | rs5943072 (3) | X | 110110885 | A | G | A |  | 0.026029 | 0.0028014 | 1.54x10<sup>-20</sup> |
| | | - | rs146191998 (1) | X | 109823511 | T | C | T | | 0.0649401 | 0.00701231 | $2.05 \times 10^{-20}$ |
| | | *BCORL1* | rs5932715 (2) | X | 129147373 | A | G | A | Missense Variant | -<br>0.0297389 | 0.00372865 | $1.53 \times 10^{-15}$ |
| | Testosterone | - | rs17307280 (11) | X | 8916646 | C | A | C | - | 0.0956476 | 0.00270341 | $4.26 \times 10^{-273}$ |
| | | - | rs1573036 (4) | X | 109820068 | T | C | T | - | 0.0659621 | 0.00245616 | $1.67 \times 10^{-158}$ |
| | | - | rs1232110 (2) | X | 8861983 | C | T | C | - | -<br>0.0411298 | 0.00311179 | $7.32 \times 10^{-40}$ |
| | | - | rs138283230 (2) | X | 8868794 | G | A | G | - | 0.083212 | 0.00744128 | $5.10 \times 10^{-29}$ |
| | | *AR* | rs147967903 (2) | X | 66874994 | G | A | G | Intron Variant | 0.103758 | 0.00937201 | $1.78 \times 10^{-28}$ |
| | DLBCL | - | rs192974431 (0) | X | 75556206 | T | C | T | - | 14.0807 | 0.429807 | $7.58 \times 10^{-10}$ |
| | | - | rs187744923 (0) | X | 95033720 | T | C | T | - | 4.82477 | 0.267175 | $3.85 \times 10^{-09}$ |
| | | *SYTL5* | rs189567150 (0) | X | 37899813 | G | A | G | Intron Variant | 6.8785 | 0.332428 | $6.59 \times 10^{-09}$ |
| | | - | rs151190138 (0) | X | 35592995 | C | G | C | - | 4.78073 | 0.270839 | $7.61 \times 10^{-09}$ |
| | | *X:1874652_C_T (0)* | | XY | 1874652 | T | C | T | - | 10.0564 | 0.408315 | $1.58 \times 10^{-08}$ |

SNPs significantly associated (p<5×10^-5^) with each lymphoma subtype were assigned to the nearest gene using the online VEP tool, generating gene profiles. Interestingly, whilst HL and NHL, as well as DLBCL and FL, have similar phenotypic presentations, our data revealed an overlap of approximately only 5 to 20% between the gene profiles of these subtypes (Fig S2). Gene profiles were also seen to be largely unique between males and females, with only up to 9.7% overlapping autosomal genes (Fig**Error! Reference source not found.** S1 A-E), and 12.9% overlapping sex chromosome encoded genes (Fig**Error! Reference source not found.** S1 F-J). Females presented a higher proportion of unique genes (ranging from 48.7 to 70.2%) compared to males (23.7 to 38.4%) (Fig S1**Error! Reference source not found.**).

Given the sex differences observed in these lymphoma subtype comparisons, a series of sex-stratified GWA analyses were performed to identify genetic variants significantly associated with sex-related characteristics, namely, oestrogen, testosterone and sex hormone binding globulin (SHBG) circulating hormone levels. The top five most significant associations, in autosomal or sex chromosome genes, are summarised in Table 3, and Table 4 respectively. Previously published associations with these significantly associated genes and haematological malignancies are summarised in S6**Table 4**.

**Table 4.**
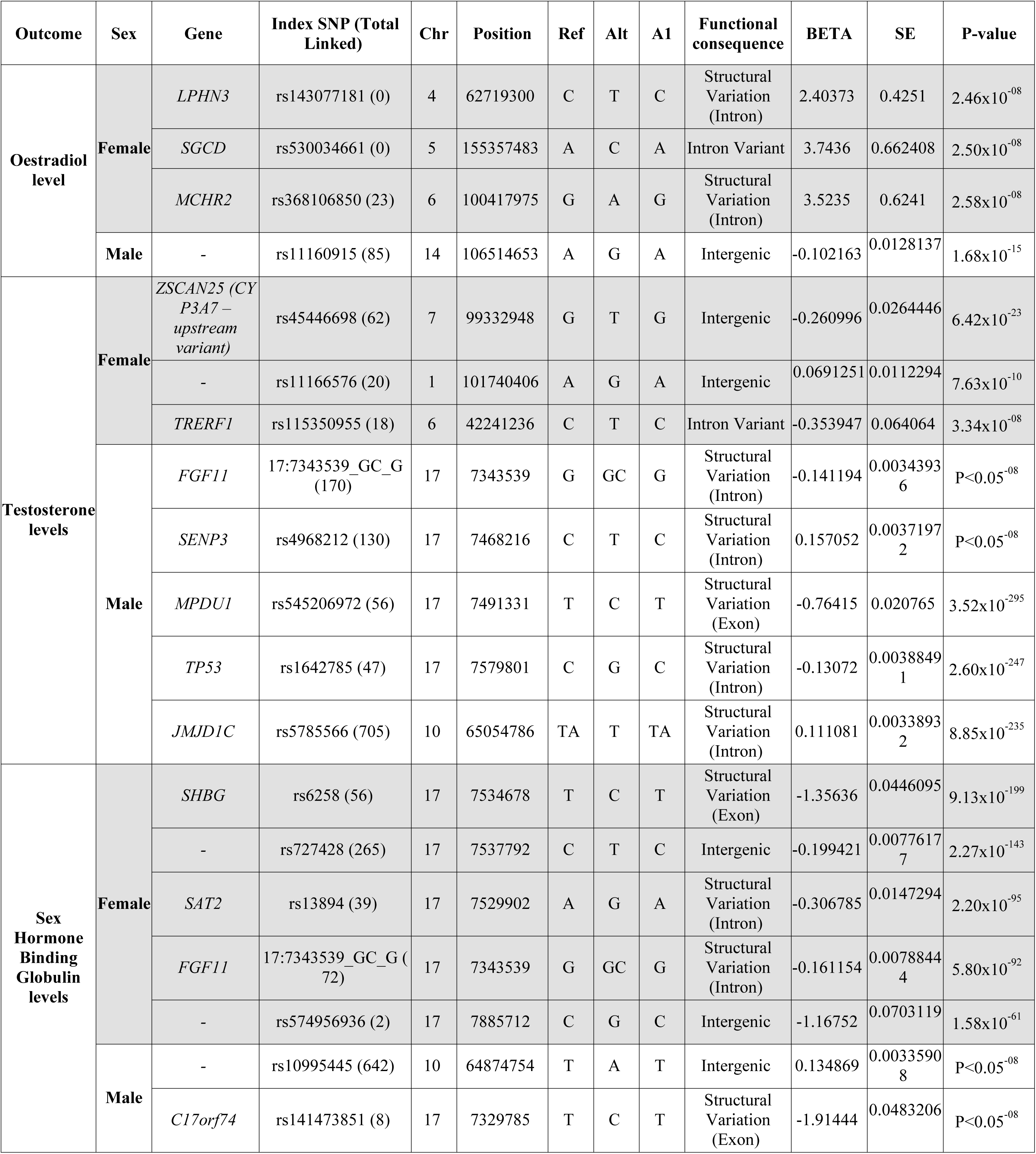
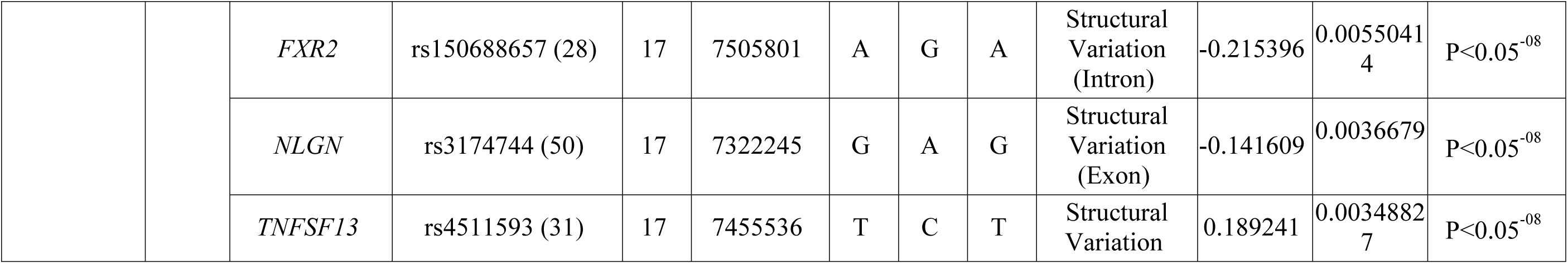
SNPs associated with hormone level determination in the UK Biobank cohort according to sex. Abbreviations: A1: affect allele, Alt: alternate allele, Chr: chromosome, Ref: reference allele, SE standard error, SNP: single nucleotide polymorphism.

As before, SNPs significantly associated (p<5×10^-5^) with each sex-related characteristic were assigned to the nearest gene using the online VEP tool, generating gene profiles. A small proportion of overlap (ranging from 0 to 7.8%) was observed between males and females for each sex-related characteristic (Fig S3). Comparing the gene profiles for each hormone, stratified by sex, revealed only a modest overlap, except for the male testosterone and SHBG gene profiles (20.7% overlapping autosomal genes, 13.9% overlapping sex chromosome encoded genes) (Fig S4**Error! Reference source not found.**). These results highlight the presence of distinct gene profiles, and therefore probable distinct cellular pathways, associated with oestrogen, testosterone and SHBG circulating hormone levels in males and females.

Following this, we investigated the intersection between genes containing genetic variants significantly associated with each lymphoma subtype, and those genes containing genetic variants significantly associated with circulating hormone levels. Interestingly, the highest proportion of overlap was observed between lymphoma subtypes and oestrogen in females (3.5 to 4.3% in autosomal genes (Fig S5 A-E), 9.1% in sex chromosome encoded genes (Fig S5 F-J)). In males, only modest overlap (ranging from 0.7 to 3.1%) was observed (Fig S6).

To gain functional insights into the shared underlying biology between the pathways associated with lymphoma and hormone level determination, gene ontology analysis was undertaken. Gene profiles, stratified by sex, were evaluated against the complete human genetic background to identify enriched biological processes, using the TopGO *classic* algorithm [31].

Harnessing the gene ontology clustering offered by the ViSEAGO R package [28], functional profiles were compared via heatmaps (Fig 2). Summarised terms for each gene ontology cluster are displayed in S7. Interesting overlaps were observed between NHL and HL, or DLBCL and FL, highlighting common biological processes which may become dysregulated in these closely related subtypes. Interestingly, the JAK-STAT pathway, which has previously been associated with lymphoid malignancies [33], was significantly enriched when assessing DLBCL in males only (p-value=1.37×10^-4^, Tables S7 and S13). Functional overlaps were also observed between lymphoma subtypes and oestrogen level determination in females, as well as testosterone or SHBG level determination in males. Similar results were observed when analysing sex chromosome encoded genes (Fig S10, S8), or when harnessing the more conservative TopGO elim algorithm (Fig S11, S12, S13and S9S10) [34].

**Fig 2.**
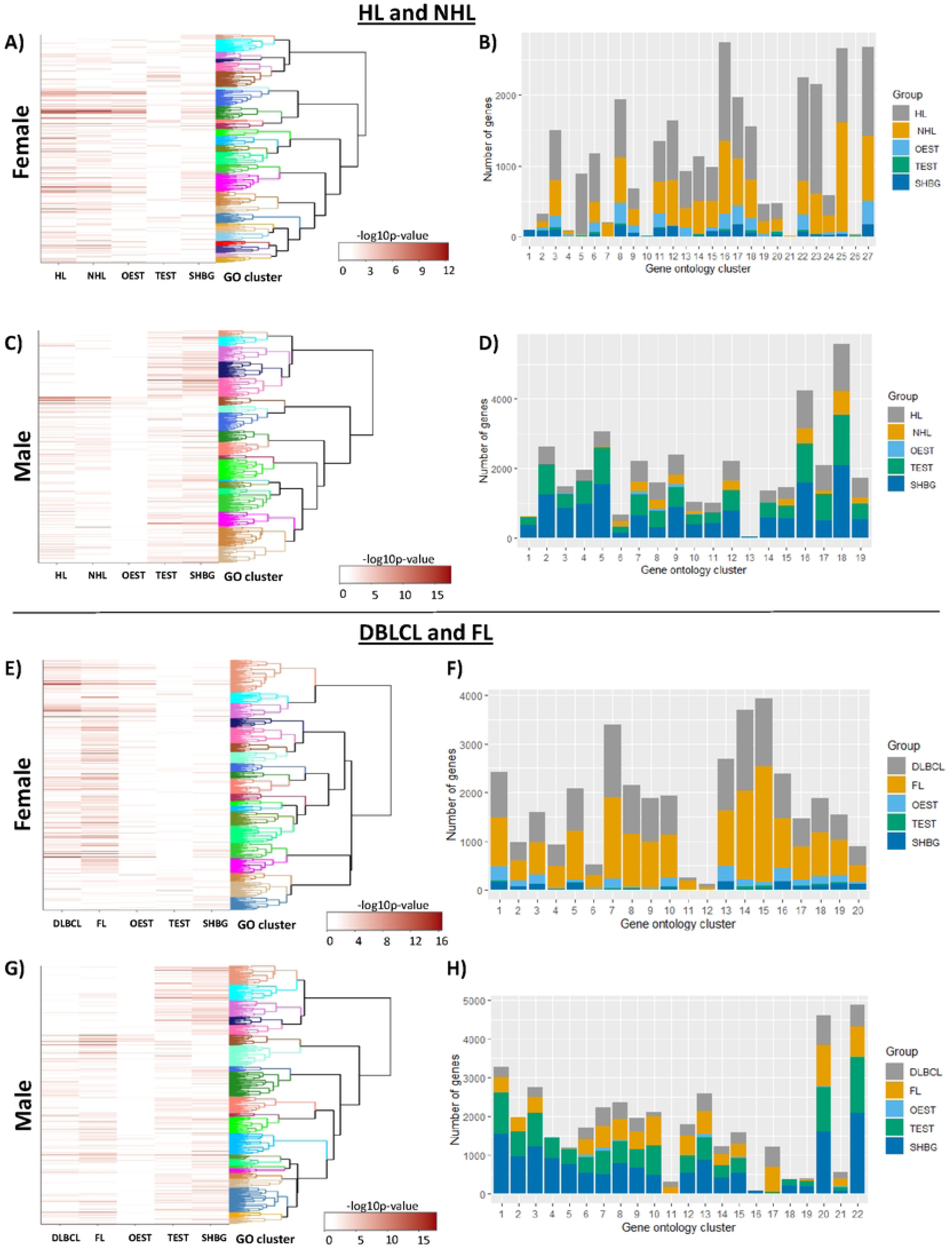
Overlapping gene ontologies for autosomal genes associated with lymphoma subtypes or hormone level determination. Upset plots representing overlaps between enriched GO terms associated with oestrogen (OEST), testosterone (TEST) and SHBG hormone levels, together with HL and NHL (A - D), or DLBCL and Follicular lymphoma (FL) (E-H). Data generated using autosomal genes (A, B, E, F) or sex chromosomes encoded genes (C, D, G, H), stratified into female (A, C E, G) or male (B, D, F, H) participants.

This gene ontology analysis highlighted terms such as nervous system development, cell communication, cell projection organisation, epigenetic modification, gene expression regulation, and cell signalling shared between lymphoma subtype pathogenesis and oestrogen level determination in females. Processes such as metabolic regulation, synapse organisation, adhesion, and behaviour were overlapping between lymphoma subtype pathogenesis and testosterone level determination in males, highlighting interesting gene and biological pathway targets for the future study of lymphoma pathogenesis sex-specificities. Upset plots shown in Fig 3 also reflect these overlaps; however, they also highlight that lymphoma subtypes and sex-related characteristics also presented unique gene ontology profiles.

**Fig 3.**
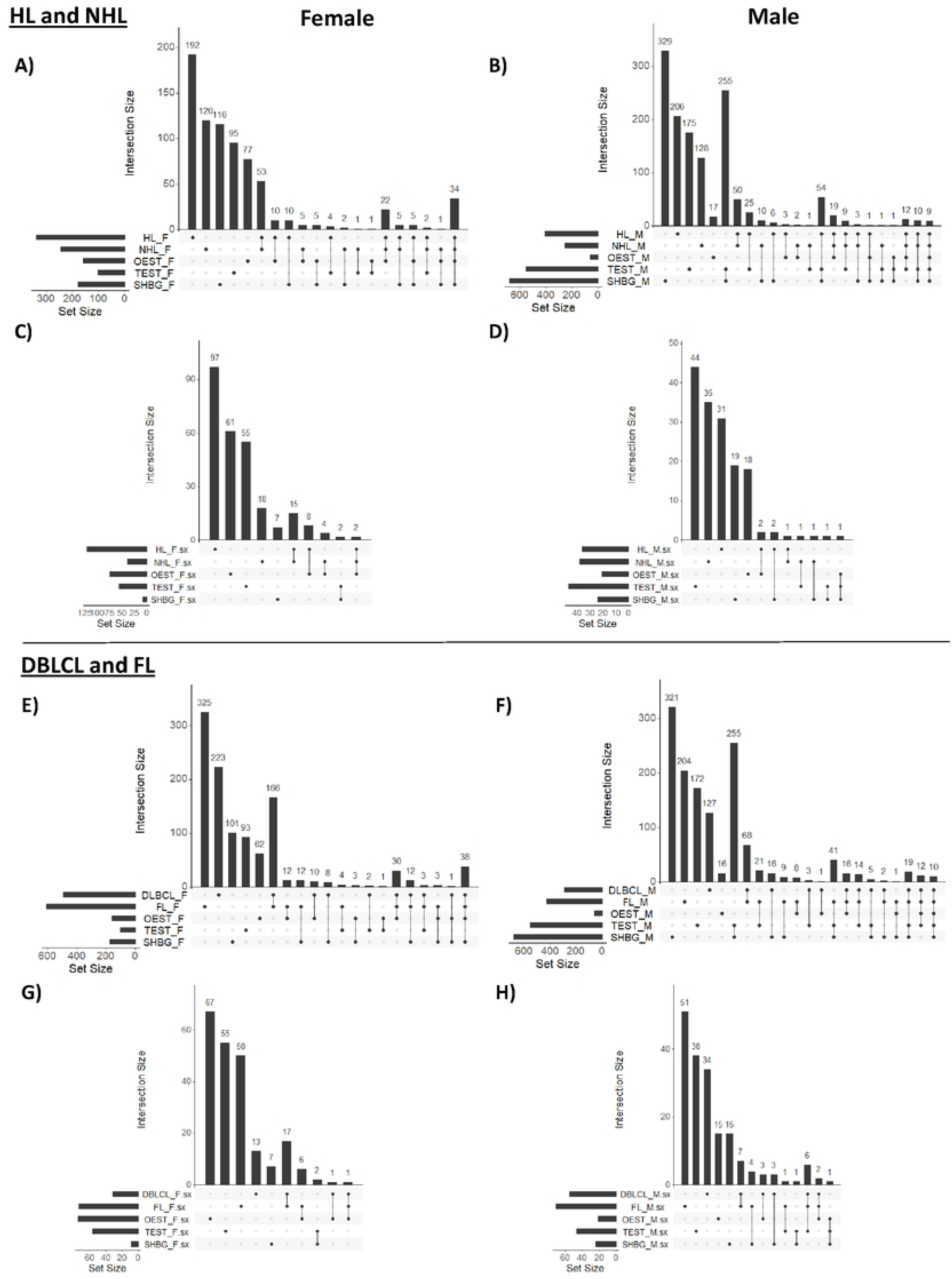
Gene ontology clustering for gene ontologies associated with lymphoma subtypes or hormone level determination. Gene ontology cluster profiles for as oestrogen (OEST), testosterone (TEST) and SHGB hormone levels, together with NL and NHL (A - D), or DLBCL and Follicular lymphoma (FL) (E-H) for autosomal genes. Heatmaps compare biological process gene ontology (GO) terms enriched for each outcome (A, C, E, G), with bar graphs depicting the number of genes present within each gene ontology cluster (B, D, F, H), from top to bottom in the corresponding heatmap, stratified into female (A, B E, F) or male (C, D, G, H) participants. Summarised terms for these gene ontology clusters are shown in S7. Raw data is shown in Table S12-16.

## Discussion

Harnessing the UKB, we explored sex differences in the development of lymphoma across a European population. For each lymphoma subtype studied (HL, NHL, DLBCL and FL), a higher disease incidence and prevalence was observed in males compared to females, concordant with previous studies highlighting higher levels of cancer, including haematological malignancies, in males [5–8,35–39].

Due to the observed sex differences in lymphoma cases, it was important for this study to incorporate sex chromosome GWA analyses. The majority of GWA studies omit chromosome X or Y analysis, potentially due to lower genotyping accuracy for chromosome X [40], or assumptions that chromosome Y is a “genetic wasteland” [41,42]. Despite this, some studies have defined the importance of including the X chromosome in GWA analyses, by highlighting variants located on the X chromosome that are associated with factors such as height, fasting insulin levels, Parkinson’s disease and infectious disease susceptibility [43–45]. Conde *et al.* explored X chromosome genetic variants associated with FL in a meta-analysis of studies with modest sample size, revealing 8 nominally associated (FDR < 0.05) independent loci (121 SNPs) [46]. Five of these SNPs were present within this sex-stratified UKB analysis; however, these SNPs were not significantly associated with FL, in either males (0.199 ≤ P ≤ 0.797) or females (0.351 ≤ P ≤ 0.874) (S11). Significantly associated X chromosome genetic variants were identified for NHL and DLCBL in this study, for example, variants within the genes *SYTL5* (in males) and *PPP2R3B* (in females); genes previously implicated in biological processes associated with haematological malignancy [47–49]. Additional genetic variants outside known coding regions of the X chromosome were also identified, with future work needed to explore the function of these genetic variants in processes such as gene regulation. Interestingly, no associations were found on the Y chromosome in this study. Challenges remain in the study of Y chromosome associations, such as poor imputation, lack of statistical power and underrepresentation in genotyping arrays [50,51].

Beyond the inclusion of sex chromosome analysis, we also performed sex-stratified analyses. Males and females had relatively unique genetic variant gene profiles, suggesting that different cellular processes may exist in males and females which influence lymphoma development. This study provided insights into the biological processes potentially overlapping between hormone level determination and lymphoma pathogenesis. Noticeably, the largest degree of overlap was observed in females between lymphoma subtypes and oestrogen level determination, for both autosomal and sex-chromosome encoded genes. There is evidence of a protective effect of oestrogens in lymphoma development. A poorer prognosis and survival rate in men versus women is predominantly seen before women enter menopause [52]. An association has also been described between decreased risk of NHL and hormone replacement therapy, contraceptive use or pregnancy, highlighting the potential protective effect of female sex hormones [53–55]. Immune function variation has been linked to circulating sex steroid hormone concentrations, interacting with genetic, epigenetic and/or environmental factors to influence the immune response [13,56], with many immune related genes having oestrogen response elements in their promotor regions [57].

Oestrogens have a prominent effect on the immune system and have also been implicated in lymphoma growth *in vitro* and *in vivo* [58,59]. Whilst oestrogen receptor alpha (ERα) and oestrogen receptor beta (ERβ) are both expressed in a range of immune cells [58], ERβ is the predominant ER in mature lymphocytes and in haematological malignancies including HL, DLBCL, Burkitt lymphoma, and multiple myeloma [59–61]. Oestrogens have been shown to promote proliferation via ERα [62,63], or supress proliferation and stimulate differentiation via ERβ [58,64,65]. Shim *et al.* highlighted that a balance between ERα and ERβ was important in haematopoiesis in bone marrow [66]. ERβ has been identified as a prognostic biomarker for DLBCL patients, with higher nuclear ERβ1 significantly associated with favourable prognosis [67]. ERα and ERβ are already useful prognosis markers for breast cancer [68–70], and interestingly, ERβ has recently been identified as a tamoxifen (a common breast cancer therapeutic)-sensitive target for DLBCL [61].

Suppression of lymphoma growth has been achieved in a range of mouse models (for both murine and human grafted lymphoma) via treatment with the ERβ agonist diarylpropionitrile (DPN) [67,71], with authors highlighting that although this was achieved in several lymphoma subtypes, the precise mechanisms may be subtype specific [72]. This is reflected in our analysis, whereby approximately one third of genes associated with oestrogen level determination and lymphoma subtypes were shared between lymphomas which present phenotypic similarities (i.e. HL and NHL, as well as DLBCL and FL). This result suggested the presence of both shared and distinct oestrogen-related biological pathways which may become disrupted during the pathogenesis of even closely related lymphomas. Our gene ontology analysis then translated these distinct gene profiles into functional insights. Interestingly, a recent study by Huang *et al.* explored the transcriptome and chromatin binding profile changes which occurred after DPN treatment in a Mantle Cell Lymphoma Tumor Model. Notably, a number of the signalling pathways affected by DPN (i.e. Wnt signalling and cell junction organisation) [71] were also highlighted in this study as functions shared between oestrogen level determination and lymphoma subtype pathogenesis. Our study has identified potentially useful pathways to aid diagnosis and advance the development of subtype specific therapies. Targeting the oestrogen pathway may be particularly useful if anti-oestrogen therapies for other cancers could be repurposed for the treatment of lymphoma.

Also relevant to the potential influence of oestrogen on lymphoma development in this study was the identification of a variant in female NHL patients (6:152322665_GT_G, risk allele G) which mapped to *ESR1*, the gene coding for ERα. ERα was related to haematological malignancies as early as 1996, when it was shown that ERα was aberrantly methylated in 86% of all hematopoietic neoplasms [73]. ERα methylation (ERM) is known to repress *ERα* gene transcription in acute myeloid leukaemia (AML), highlighting the relevance of oestrogen regulation for other haematological malignancy subtypes [74]. However, conflicting reports on whether ERM results in improved or worse outcomes has made the functional consequence of this methylation difficult to define [74–77]. Via GWA analysis, we have demonstrated an additional layer of genomic regulation, single nucleotide variation within the *ESR1* gene, which is potentially involved in NHL pathogenesis in females. Espín-Pérez *et al.* recently identified sites of differential methylation significantly associated with future development of NHL, with seven of these sites associated with the *ESR1* gene (cg00655307, cg05171584, cg07059469, cg10441070, cg13612689, cg15980539, cg25565730) [78]. The same can be said for *ALCAM* and *BIR6*, which we found to contain genetic variants significantly associated with NHL, and Espín-Pérez *et al.* identified these genes as differentially methylation prior to NHL onset (*ALCAM*: cg08519233, cg09025274, cg09222287; *BIR6*: cg12922032, cg18891308, cg20351608) [78]. Given the prior evidence of interplay between genetic and epigenetic variation, resulting in allele specific gene expression in immune related diseases and cancer [79] scope exists to further investigate the interplay between genetic and epigenetic methylation of ERs in lymphoma more specifically, with potential wider implications of these interactions in the pathogenesis of other haematological malignancies.

Whist we have explored methylation data from other studies, a limitation of this study was that no exploration of epigenetic data was carried out in this cohort; due to no epigenetic data being available within the UK Biobank for this purpose at the time of analysis. An additional limitation of study was the use of a single Caucasian cohort. Future analyses to replicate these findings, particularly in an ethnically diverse cohort, would potentially further support these findings. Due to the low number of non-Caucasian ethnicities in the UK Biobank, this was not completed as a part of this analysis.

## Conclusion

Via subtype- and sex-stratified GWA analyses of lymphoma subtypes in a UKB cohort, the autosomal and sex chromosome genetic landscapes for each subtype could be compared between males and females. In addition to identifying new sex-specific genetic variants associated with specific lymphoma subtypes, we revealed that between males and females, only up to 13% similarity is observed in the genes containing genetic variants significantly associated with each lymphoma subtype. This genetic data reflects the sex differences observed in lymphoma case rates.

Previous evidence supports the influence of sex-related characteristics on haematological malignancy pathogenesis and therefore, this study explored the intersection between the genes potentially implicated in lymphoma subtypes and those involved in hormone level determination, in males and female separately. The largest overlap (up to 9.1%) was observed between lymphoma subtypes and oestrogen gene profiles in females, reflecting the known associations between oestrogen immune regulation and its potential protective effect for preventing haematological malignancy pathogenesis. Indeed, in female NHL patients, we identified a genetic variant mapping to *ESR1* (coding for oestrogen receptor-α), highlighting the potential presence of an additional layer of regulation, genetic variation, which coincides with previous observations of epigenetic *ESR1* dysregulation in NHL [78]. Gene ontology analysis also provided functional overlaps between biological processes associated with hormone regulation and lymphoma pathogeneses, for example, Wnt signalling, cell junction organisation, epigenetic modification and gene expression regulation, highlighting similarities and differences useful for guiding downstream analysis identifying novel targets for the development of more personalised therapeutic and diagnostic tools.

## Data Availability

All data is available from UK biobank upon request. Association data is available from UK biobank or from the authors upon request.

https://www.ukbiobank.ac.uk/

## List of Abbreviations

A1: Affect allele
AML: Acute myeloid leukaemia
Alt: Alternate allele
BMI: Body mass index
Chr: Chromosome
DLBCL: Diffuse large B-cell lymphoma
DPN: Diarylpropionitrile
ERα: Oestrogen receptor alpha
ERβ: Oestrogen receptor beta
F: Female
FL: Follicular lymphoma
GWA: Genome wide association
HL: Hodgkin’s lymphoma
HRT: Hormone replacement therapy
M: Male
MCL: Mantle cell lymphoma
NHL: Non-Hodgkin’s lymphoma
OC: Oral contraceptive
OEST: Oestrogen
Ref: Reference allele
SE: Standard error
SHBG: Sex hormone binding globulin
SNP: Single nucleotide polymorphism
Sx: Sex
TEST: Testosterone
UKB: UK Biobank
WES: Whole exome sequencing

## Acknowledgements

This research has been conducted using the UK Biobank Resource under Application Number 21727. Figure 1 was created with Biorender.com.

## Availability of Data and Materials

The primary data used and/or analysed during the current study are available from the UK Biobank. Detailed phenotyping information and summative data are presented in this paper.

## Author Contributions

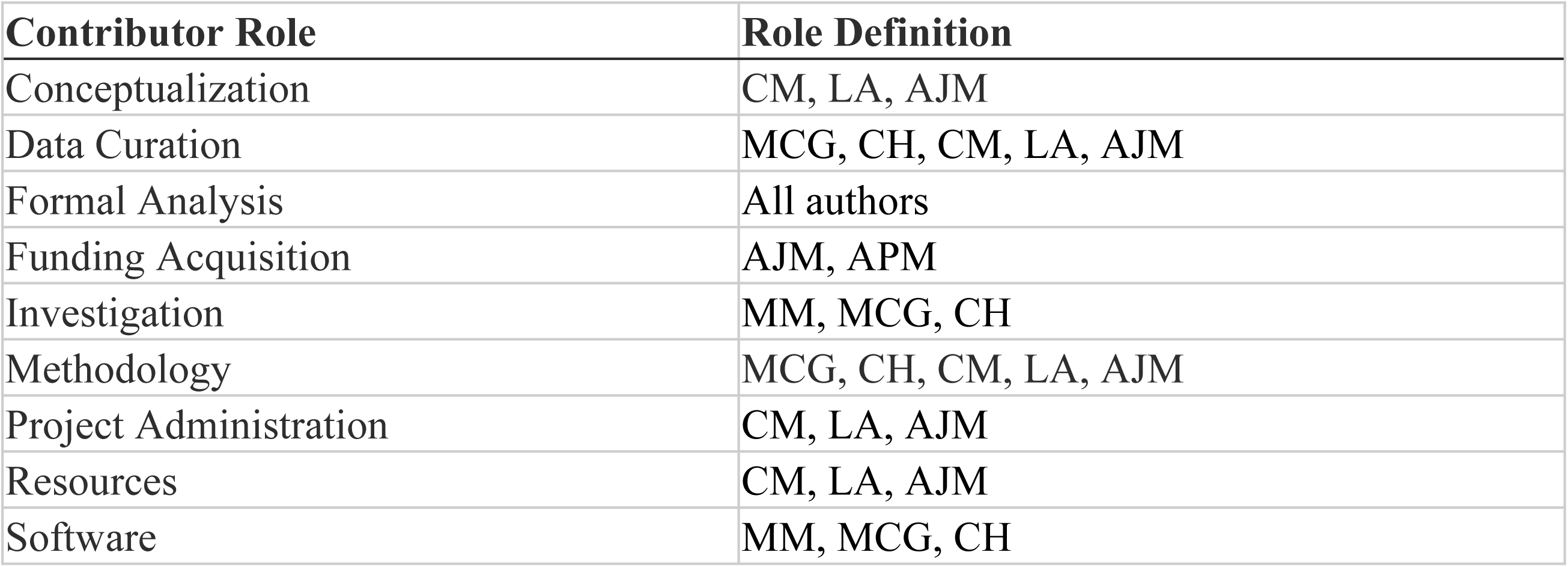
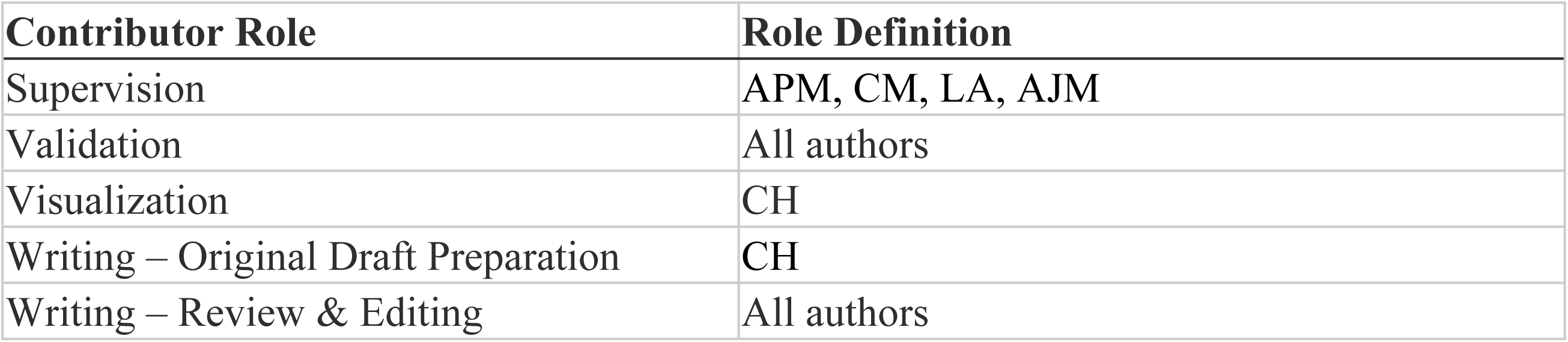

## Supporting Information

**S1 Fig. Sex comparison of autosomal or sex chromosome genes containing genetic variants significantly associated with lymphoma.** Comparison between females (F) and males (M) of autosomal (A-E) or sex chromosome encoded (F-J) genes which contain genetic variants significantly associated with each of the lymphoma subtypes investigated: All lymphomas (A, F), HL (B, G), NHL (C, H), DLBCL (D, I) and FL (E, J). Values displayed represent a percentage of the total number of genes associated (in males, females or both) with that particular lymphoma subtype. The total number of genes associated with each lymphoma subtype is displayed within each sub-figure legend.

**S2 Fig. Comparison of genes containing genetic variants significantly associated with lymphoma subtypes.** Comparison of genes containing genetic variants significantly associated with the HL and NHL (A-D) or DLBCL and FL (E-H) subgroups for females (A, C, E, G) and males (C, D, F, H), as well as autosomal (A, B, E, F) or sex chromosome (C, D, G, H) encoded genes. Values displayed represent a percentage of the total number of genes associated with those particular lymphoma outcomes, as displayed within each sub-figure legend.

**S3 Fig. Sex comparison of genes containing genetic variants significantly associated with hormone level determination.** Comparison of autosomal (A, B, C) or sex chromosome (D, E, F) encoded genes in females (F) and males (M) which contain genetic variants significantly associated with the oestrogen (Oest, A, D), testosterone (TEST, B, E) or SHBG (C, F) circulating hormone levels. Values displayed represent a percentage of the total number of genes associated (in males, females or both) with that particular hormone level outcome. The total number of genes associated with each lymphoma is displayed within each sub-figure legend.

**S4 Fig. Comparison of genes which containing genetic variants significantly associated with hormone level determination.** Comparison of autosomal (A) or sex chromosome (B) encoded genes in females (F) and males (M) which contain genetic variants significantly associated with the oestrogen (OEST), testosterone (TEST) or SHBG circulating hormone levels. Values displayed represent a percentage of the total number of genes associated (in males, females or both) with that particular hormone level outcome, as displayed within each sub-figure legend.

**S5 Fig. Comparison between genes containing genetic variants significantly associated with oestrogen, testosterone, SHBG and lymphoma subtypes (females).** Comparison between female autosomal (A-E) or sex chromosome encoded (F-J) genes which contain genetic variants significantly associated with oestrogen (OEST), testosterone (TEST), SHBG and each of the lymphoma subtypes investigated: All lymphomas (A, F), HL (B, G), NHL (C, H), DLBCL (D, I) and FL (E, J). Values displayed represent a percentage of the total number of genes associated with those outcomes, as displayed within each sub-figure legend.

**S6 Fig. Comparison between genes containing genetic variants significantly associated with oestrogen, testosterone, SHBG and lymphoma (males).** Comparison between male autosomal (A-E) or sex chromosome encoded (F-J) genes which contain genetic variants significantly associated with oestrogen (OEST), testosterone (TEST), SHBG and each of the lymphoma groups investigated: All lymphomas (A, F), HL (B, G), NHL (C, H), DLBCL (D, I) and FL (E, J). Values displayed represent a percentage of the total number of genes associated with those outcomes, as displayed within each sub-figure legend.

**S7 Fig. Overlap between lymphoma subtypes for genes significantly associated with oestrogen levels and lymphoma subtypes (females).** Comparison of autosomal (A, B) or sex chromosome (C, D) encoded genes in females which contain genetic variants significantly associated with the oestrogen (Oest) circulating hormone levels and either HL versus NHL (A, C) or DLBCL versus FL (B, D). Values displayed represent a percentage of the total number of genes, as displayed within each sub-figure legend.

**S8 Fig. Overlap between lymphoma subtypes for genes significantly associated with testosterone levels and lymphoma subtypes (males).** Comparison of autosomal (A, B) or sex chromosome (C, D) encoded genes in males which contain genetic variants significantly associated with the testosterone (Test) circulating hormone levels and either HL versus NHL (A, C) or DLBCL versus FL(B, D). Values displayed represent a percentage of the total number of genes, as displayed within each sub-figure legend.

**S9 Fig. Overlap between lymphoma subtypes for genes significantly associated with SHBG levels and lymphoma subtypes (males).** Comparison of autosomal (A, B) or sex chromosome (C, D) encoded genes in males which contain genetic variants significantly associated with the SHBG circulating hormone levels and either HL versus NHL (A, C) or DLBCL versus FL (B, D). Values displayed represent a percentage of the total number of genes, as displayed within each sub-figure legend.

**S10 Fig. Gene ontology clustering for sex chromosome genes associated with lymphoma subtypes or hormone level determination.** Gene ontology cluster profiles for as oestrogen (OEST), testosterone (TEST) and SHBG hormone levels, together with NL and NHL (A - D), or DLBCL and FL (E-H) for sex chromosome encoded genes. Heatmaps compare biological process gene ontology (GO) terms enriched for each outcome (A, C, E, G), with bar graphs depicting the number of genes present within each gene ontology cluster (B, D, F, H), from top to bottom in the corresponding heatmap, stratified into female (A, B E, F) or male (C, D, G, H) participants. Summarised terms for these gene ontology clusters are shown in S8. Raw data is shown in Table S16-19.

**S11 Fig. Gene ontology clustering for gene ontologies (elim method) associated with lymphoma subtypes or hormone level determination.** Gene ontology cluster profiles (*elim method*) for as oestrogen (OEST), testosterone (TEST) and SHGB hormone levels, together with NL and NHL (**A - D**), or DLBCL and FL (**E-H**) for autosomal genes. Heatmaps compare biological process gene ontology (GO) terms enriched for each outcome (**A, C, E, G**), with bar graphs depicting the number of genes present within each gene ontology cluster (**B, D, F, H**), from top to bottom in the corresponding heatmap, stratified into female (**A, B E, F**) or male (**C, D, G, H**) participants. Summarised terms for these gene ontology clusters are shown in **S9.**

**S1 Fig. Gene ontology clustering for sex chromosome genes (elim method) associated with lymphoma subtypes or hormone level determination**. Gene ontology cluster profiles (*elim method*) for as oestrogen (OEST), testosterone (TEST) and SHBG hormone levels, together with NL and NHL (A - D), or DLBCL and FL (E-H) for sex chromosome encoded genes. Heatmaps compare biological process gene ontology (GO) terms enriched for each outcome (A, C, E, G), with bar graphs depicting the number of genes present within each gene ontology cluster (B, D, F, H), from top to bottom in the corresponding heatmap, stratified into female (A, B E, F) or male (C, D, G, H) participants. Summarised terms for these gene ontology clusters are shown in S10.

**S13 Fig. Overlapping gene ontologies (elim method) between genes associated with lymphoma subtypes or hormone level determination.** Upset plots representing overlaps between enriched GO terms (elim method) associated with oestrogen (OEST), testosterone (TEST) and SHBG hormone levels, together with HL and NHL (A - D), or DLBCL and FL (E-H). Data generated using autosomal genes (A, B, E, F) or sex chromosomes encoded genes (C, D, G, H), stratified into female (A, C E, G) or male (B, D, F, H) participants.

**S1 Table. Definition of the outcome variables.**

**S2 Table. Definition of the Sociodemographic and clinical variables. S3 Table. Summary of PLINK quality control statistics.**

**S4 Table. Genetic variants significantly associated with lymphoma subtypes and previous associations with the development or progression of haematological malignancies.**

**S5 Table. Sex chromosome encoded genetic variants significantly associated with serum hormone level or lymphoma and previous associations with the development or progression of haematological malignancies.**

**S6 Table. Genes containing genetic variants significantly associated with serum hormone levels and previous associations with the development or progression of haematological malignancies.**

**S7 Table. Summary of gene ontology terms as shown in Fig 3 (autosomes, *classic* method)**. For gene ontology comparisons between HL and NHL (A) or DLBCL and FL (B), alongside oestrogen, testosterone and SHBG. These gene ontology terms have been manually categorised into more general categories (C) to aid comparisons.

**S8 Table. Summary of gene ontology terms as shown in Error! Reference source not found. (sex chromosomes, *classic* method)**. For gene ontology comparisons between HL and NHL (A) or DLBCL and FL (B), alongside oestrogen, testosterone and SHBG. These gene ontology terms have been manually categorised into more general categories (C) to aid comparisons.

**S9 Table. Summary of gene ontology terms as shown in S (autosomes, *elim* method)**. For gene ontology comparisons between HL and NHL (A) or DLBCL and FL (B), alongside oestrogen, testosterone and SHBG.

S10 Table. Summary of gene ontology terms as shown in Error! Reference source not found. (sex chromosomes, *elim* method). For gene ontology comparisons between HL and NHL (A) or DLBCL and FL (B), alongside oestrogen, testosterone and SHBG.

**S11 Table. SNPs from Conde et al. overlapping with the Follicular lymphoma GWAS carried out in this analysis.** None of the 5 SNPS present are significantly associated.

**S12 Table. Gene ontology clusters (and significant genes within those clusters) identified in female autosomes via the ViSEAGO classic method.** For DLBCL and FL, alongside oestrogen, testosterone and SHBG.

**S13 Table. Gene ontology clusters (and significant genes within those clusters) identified in male autosomes via the ViSEAGO classic method.** For DLBCL and FL, alongside oestrogen, testosterone and SHBG.

**S14 Table. Gene ontology clusters (and significant genes within those clusters) identified in female autosomes via the ViSEAGO classic method.** For NHL and HL, alongside oestrogen, testosterone and SHBG.

**S15 Table. Gene ontology clusters (and significant genes within those clusters) identified in male autosomes via the ViSEAGO classic method.** For NHL and HL, alongside oestrogen, testosterone and SHBG.

**S16 Table. Gene ontology clusters (and significant genes within those clusters) identified in female sex chromosome via the ViSEAGO classic method.** For DLBCL and FL, alongside oestrogen, testosterone and SHBG.

**S17 Table. Gene ontology clusters (and significant genes within those clusters) identified in male sex chromosome via the ViSEAGO classic method.** For DLBCL and FL, alongside oestrogen, testosterone and SHBG.

**S18 Table. Gene ontology clusters (and significant genes within those clusters) identified in female sex chromosomes via the ViSEAGO classic method.** For NHL and HL, alongside oestrogen, testosterone and SHBG.

**S19 Table. Gene ontology clusters (and significant genes within those clusters) identified in male sex chromosomes via the ViSEAGO classic method.** For NHL and HL, alongside oestrogen, testosterone and SHBG.

